# ChatGPT with Mixed-Integer Linear Programming for Precision Nutrition Recommendations

**DOI:** 10.64898/2026.02.14.26346312

**Authors:** Rozalina Alkeyeva, Ilyas Nagiyev, Damir Kim, Bibinur Nurmanova, Zhuldyz Omarova, Huseyin Atakan Varol, Mei-Yen Chan

**Affiliations:** School of Sciences and Humanities, Nazarbayev University, Astana 010000, Kazakhstan; School of Medicine, Nazarbayev University, Astana 010000, Kazakhstan; Institute of Smart Systems and Artificial Intelligence, Nazarbayev University, Astana 010000, Kazakhstan

**Author notes:** Correspondence (M.-Y.C.). These authors contributed equally to this work (Rozalina Alkeyeva, Ilyas Nagiyev, Damir Kim). (R. A.); (I. N.); (D.K.); (B. N.); (Z. O.).

**Keywords:** Artificial intelligence, Large language model (LLM), AI-powered nutrition chatbot, Personalized nutrition, Chatbot, Dietary assessment, Validation

## Abstract

**Background:** The growing interest in applying artificial intelligence in personalized nutrition is challenged by the complex nature of dietary advice that must balance health, economic, and personal factors. Though automated solutions using either Linear Programming (LP) or Large Language Models (LLMs) already exist, they have significant drawbacks. LP often lacks personalization, whereas LLMs can be unreliable for precise calculations.

**Objectives:** To develop and assess a model that integrates a Mixed Integer Linear Programming (MILP) solver with an LLM to generate personalized meal plans and compare it with standalone LLM and MILP models.

**Methods:** The proposed hybrid MILP+LLM model first uses an LLM (GPT-4o) to filter a unified food dataset (n=297), which combines regional Central Asian and global food items, according to the user’s profile. The filtered list of food items is then received by a MILP solver which identifies the set of top 10 optimal solutions. Finally, given this set of solutions, LLM chooses the most appropriate meal plan. The model was evaluated using five synthesized, clinically complex patient profiles sourced from Adilmetova et al. [4]. The performance of this hybrid model was compared against standalone MILP and LLM using 5-point Likert scale with Kruskal-Wallis and post hoc Dunn’s tests for Nutrient Accuracy, Personalization, Practicality, and Variety.

**Results:** Findings demonstrated that the proposed MILP+LLM model reached balanced performance achieving scores of more than 3.6 points in all criteria, with high scores in Nutrient Accuracy (3.96), Personalization (3.81), and Practicality (3.99). The standalone LLM model performed the weakest in all criteria, with statistically significant lower scores compared to the other two methods. The standalone MILP model performed best in Nutrient Accuracy (4.93) and in Variety (4.10) but lagged behind the MILP+LLM model in Practicality and Personalization. Kruskal-Wallis and Dunn’s tests showed MILP and MILP+LLM outperformed LLM across all criteria. MILP was more accurate (p<0.0001), while MILP+LLM model was more practical (p=0.021).

**Conclusions:** The findings suggest that integrating the LLM with the MILP solver creates a model that combines qualitative personalization with quantitative precision. This model produces comprehensive, reliable meal plans, addressing the limitations of using either model alone.

## 1. Introduction

Research on personalized nutrition has emphasized the increasing importance of tailoring dietary interventions to individual needs. This personalized approach, when compared to generalized dietary guidelines, has been observed to yield better health outcomes and improved adherence among patients [1,2]. However, the design of personalized plans frequently depends on a wide range of dynamic factors, making it a complex problem. These factors include patients’ unique health conditions, physiological needs, cultural norms, personal taste preferences, economic constraints, and the seasonal availability of ingredients. It is crucial to overcome this challenge, especially considering the direct relation between poor dietary habits and the development of chronic health conditions such as type 2 diabetes, hypertension, cardiovascular diseases, obesity, and certain types of cancer [1].

Manually designing personalized meal plans that account for the aforementioned factors can be challenging and time-consuming for individuals, as well as for human nutrition experts [3]. Although traditional nutritional counseling remains an invaluable source of dietary guidance, there are substantial access barriers caused by the high costs, impracticality, and non-availability to the broader population [3, 4]. These constraints on human expertise highlight the inefficiency of manual dietary design approaches in simultaneously optimizing complex and dynamic dietary variables such as health conditions, taste, and budget. Therefore, there is a growing need to develop sophisticated automated solutions that can process and integrate this significant amount of data to provide people with scalable and accessible personalized nutrition.

Linear programming (LP) has been the subject of interest for many studies in diet optimization, with its origins starting from George Stigler’s seminal “diet problem” in the 1940s [5]. This work pioneered the transformation of complex nutritional challenges into solvable mathematical models. In nutrition, these models are referred to as a diet optimization models which, given certain set of constraints (e.g., nutritional requirements and amounts of food), aim to find optimal combination of food items that minimizes or maximizes an objective function (e.g., total deviation from nutrient goals and the total cost of diet) [6]. LP, as one such method, offers a robust mathematical framework for solving diet optimization problems. Over the last several decades, a number of attempts have been made to utilize LP for generating dietary recommendations. Among prominent examples, there are World Health Organization’s Optifood and World Food Programme’s NutVal that have successfully applied LP to design sustainable diets optimized for maximizing nutrient intake within realistic dietary constraints [7]. Apart from using LP to optimize diet for nutritional requirements, several studies have explored its applications for minimizing meal costs and producing budget-friendly dietary recommendations [8,9]. Although in most cases, LP models are formulated using quantitative data, previous research has also made efforts to adapt them for more nuanced applications. For instance, Salloum and Tekli [3] adapted “transportation optimization problem” (a form of LP) to develop the Meal Plan Generator (MPG) solution that accounts for factors such as food preferences and inter-food compatibility. This way, quantification of qualitative variables allowed LP to become a multi-objective optimization framework, emphasizing its importance in automated meal planning aimed at identifying optimal food combinations for complex nutritional target values.

The recent advances in Large Language Models (LLMs) to understand and generate human-like text have resulted in considerable interest around their applications to personalized nutrition. A variety of studies established that LLMs offer a promising solution to the current challenges with accessibility and practicality of personalized nutritional counseling, allowing users to receive effective dietary recommendations that, in some cases, even outperform guidance from human dietitians [1, 10, 11]. For example, Damette and colleagues introduced an LLM-based Meal Planning Assistant aimed at helping health professionals in designing personalized meal plans, which utilized GPT-4o to interpret patient profiles [10]. LLMs were also found to perform well when integrated into multi-modal systems, such as a prototype for a digital nutrition assistant where an LLM was combined with a computer vision module to generate personalized nutritional guidance tailored to a specific culinary landscape [12]. The main advantage of LLMs lies in their capability to ‘extract’ qualitative preferences and objectives (e.g., taste preferences, preferred cuisines, and allergies) from user profiles. This is a feature typically absent or difficult to integrate in rule-based optimization models. This way, due to a wide range of world knowledge and capability to engage in natural language conversations, LLMs can generate diverse and user-friendly personalized dietary recommendations, making nutritional guidance accessible to everyone.

While both LP-based and LLM-powered meal designing approaches contribute to automated food recommendation systems, each method possesses certain limitations that undermine their capabilities. Despite the ability of dietary recommendation systems based on LP as demonstrated in the MPG [3] to incorporate qualitative factors from user profiles through cost functions, they are dependent on pre-defined rules and numerical values which were used as a method of conversion of natural language inputs. As a result, LP-generated dietary suggestions may lack flexibility and adaptability to individual dynamic dietary factors [7]. LLMs, in turn, often fail to provide reliable nutritional analyses due to the absence of explicit computational algorithms in their functionality. Since LLMs fundamentally operate on statistical patterns and relationships learned from text data, they lack a deeper understanding of the underlying scientific principles in nutritional analysis. In other words, while LLMs are able to ‘talk about’ nutrition, they fail to ‘do’ nutrition in a quantifiable manner that is essential for reliable nutritional guidance. Therefore, LLMs’ inability to accurately calculate nutrient values of food items and extract information from verifiable nutritional databases poses a significant challenge to conducting precise and detailed dietary assessment. For instance, previous research underscored LLMs’ limitations in accounting for critical food components (e.g. calories, proteins, and fat) [10, 13, 14]. This is exemplified in the study conducted by Bergling et al. [13], where the evaluation of GPT-4’s accuracy in generating dietary recommendations for patients with chronic kidney disease revealed underestimation of calories, protein, and fat by 36%, 28%, and 48%, respectively. Considering these limitations of both standalone LLM and standalone LP solutions, it can be observed that the weaknesses of one approach are precisely the strengths of the other and vice versa. This presents a strong motivation for this work carried out to combine both approaches in a single hybrid model for personalized diet optimization.

The project aims to propose and evaluate a hybrid model that combines the advantages of Mixed Integer Linear Programming (MILP) solvers with LLMs for generating personalized meal plans. This model aims to assist the LLMs with accuracy in computations while enhancing the personalization of LP. To the best of the authors’ knowledge, this is the first model to integrate LLMs and MILP solvers for structured, personalized meal planning with mock patient-specific profiles.

Our work is shaped by successful implementation of hybrid AI systems integrating LLMs with optimization algorithms in other domains requiring complex personalized planning. A key example is Google Research’s hybrid trip-planning system that utilizes an LLM for generating an initial plan based on user preferences that is further refined by an optimization layer for ensuring feasibility against real-world constraints such as travel times and opening hours [15]. This system provides a strong methodological precedent for our work.

In the field of personalized nutrition, several hybrid frameworks have emerged combining LLMs with a wide range of methods such as integration with nutrition databases or recipe datasets, Retrieval-Augmented Generation (RAG), and sophisticated prompt engineering [10, 14, 16]. A prominent example is the “Multi-objective Personalized Health-aware Food Recommendation System” framework developed by Zhang et al. [17] which employs Pareto optimization module based on gradient descent algorithm to jointly optimize for user preferences, healthiness and nutritional diversity, while LLMs are used for generating explanatory reasoning for the user. However, existing studies do not explicitly state the use of mathematical programming solvers for satisfying hard quantitative constraints such as caloric limits and nutrient targets. Their optimization approaches are focused on balancing soft goals rather than providing precise numerical guarantees essential for adhering to dietary requirements. The core novelty of our approach lies in the explicit integration of an LLM with MILP solver, ensuring that generated dietary advice are not only personalized and well-explained but also compliant with complex quantitative constraints.

## 2. Methods

This section provides a detailed description of the proposed hybrid MILP+LLM model, the design of the comparative standalone models, and the evaluation methodology utilized for assessing the performance of each of three approaches.

### 2.1. Hybrid MILP+LLM Model Architecture

Our model combines a LLM and a MILP solver to create a multi-stage pipeline for personalized dietary recommendations generation. The system is provided with a list of available food items from a pre-compiled food dataset. The workflow begins with the construction of a structured prompt for LLM, incorporating the food products and a given patient-specific medical profile. The algorithm formulates the prompt using a standardized template that combines the complete food list with detailed patient medical information. The LLM analyzes the patient profile against the available food options and generates a filtered list of appropriate food items. The curated list is then forwarded to the MILP solver, which optimizes meal combinations against desired caloric and macronutrient targets. The solver identifies a set of top 10 optimal meal plans, which are subsequently passed to the LLM for final selection. At the final stage, the LLM is prompted to choose the single best plan and provide detailed rationale for its choice. For improved user interaction, an image of the proposed meal plan is generated using an AI-based image generation model. The following subsections provide a detailed description of the main components of the proposed architecture.

#### 2.1.1 Dataset Construction

To capture both global variety and regional specificity, the unified dataset was constructed using two complementary sources. Its foundation is an internal Central Asian food database, consisting of 139 dishes that are commonly consumed in the region. This database was developed based on the publicly available Central Asian Food Dataset introduced in Karabay et al. [18], as well as the more recent dataset described by Sanatbyek et al. [19,20]. To add variety and expand coverage with globally popular dishes, items were incorporated from the Food Nutrition Dataset by Utsav Dey [21], an open Kaggle collection containing over 1,000 food items. From this source, 158 items were manually selected by a team of nutritionists, aimed at increasing food variety, ensuring dietary diversity and avoiding overlaps with the Central Asian dataset. The final unified food database thus contains 297 food items.

Each food item includes a complete feature set with standardized macronutrient values (calories, protein, carbohydrates, fats, potassium, and sodium) per gram. The dataset is organized into six food categories—main dish, soup, salad, drink, snack, and dessert. Table 1 summarizes the distribution of items across these categories, with separate counts shown for the Central Asian dataset, the Utsav Dey selections, and the combined dataset.

**Table 1.**
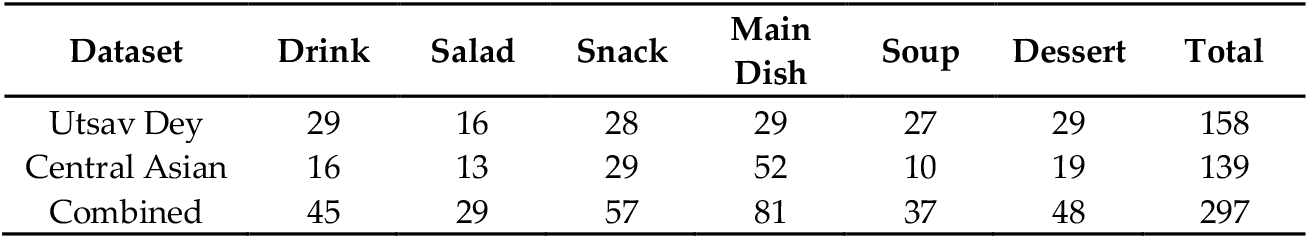
Distribution of food items from the study by category across the Central Asian, Utsav Dey, and combined datasets [18-21].

| Dataset | Drink | Salad | Snack | Main Dish | Soup | Dessert | Total |
| --- | --- | --- | --- | --- | --- | --- | --- |
| Utsav Dey | 29 | 16 | 28 | 29 | 27 | 29 | 158 |
| Central Asian | 16 | 13 | 29 | 52 | 10 | 19 | 139 |
| Combined | 45 | 29 | 57 | 81 | 37 | 48 | 297 |

#### 2.1.2 Initial Filtering by LLM

The proposed pipeline starts with a filtering process conducted using the LLM to facilitate personalization of the food item selection. Given the food dataset and a detailed patient profile (Appendix 1) consisting of a user’s demographic information, health conditions and goals, dietary habits, and preferences, the LLM is tasked with identifying and removing unsuitable food items, such as allergens, culturally inappropriate foods, and items that contradict medical advice. The main aim of this preprocessing step is to improve further optimization process by restricting the solution space to relevant options only.

The GPT-4o model was selected as the LLM for this task, as recent studies have shown that it delivers strong performance in producing coherent and relevant dietary advice [22,23]. Prompt engineering methods were utilized to ensure consistent and focused responses from the model. The system prompt was formulated to define the LLM’s role as well as constrain its output format as following:

“You are a dietitian. In your reply, output ONLY a Python list literal of the item names to KEEP.”

This instruction was combined into a single text with the patient profile information and the full list of food items from the dataset, formatted to ensure clarity and consistent parsing. The system parameters were configured to minimize response variability and ensure deterministic outputs.

#### 2.1.3 Optimization using a MILP Solver

The core optimization part of the proposed dietary recommendation system was handled by the MILP solver. For its implementation Python’s PuLP library was used. After receiving the list of food items from the initial filtering step conducted by LLM, the solver proceeds with constructing meal plans optimized for the patient’s unique nutritional requirements, specifically caloric and macronutrient targets. The MILP solver is tasked with finding the top 10 optimal meal combinations which are subsequently offered to LLM for the final qualitative selection.

To establish precise and personalized nutritional targets, the patient’s Basal Metabolic Rate (BMR) is first calculated based on their profile data (sex, height, weight, and age) using the following Mifflin-St Jeor equations [24]:

- For men:

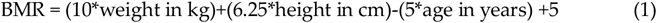
- For women:

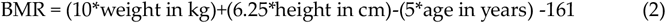

The obtained BMR value is then multiplied by the patient’s stated physical activity level to estimate Total Daily Energy Expenditure (TDEE). Table 2 provides 5 physical activity levels utilized for the calculation. Assuming three-meal daily structure, one-third of the TDEE was used as the final caloric target of the meal.

**Table 2.** Physical activity level [25].

| Physical Activity Level (PAL) | Multiplier |
| --- | --- |
| Sedentary (little to no exercise) | 1.2 |
| Lightly active (1-3 days/week) | 1.375 |
| Moderately active (3-5 days/week) | 1.55 |
| Very active (6-days/week) | 1.725 |
| Extra active (athletes, physical jobs) | 1.9 |

The total calorie goal obtained from the patient’s input data is then distributed across macronutrients according to the following target percentages suggested by Dietary Guidelines for Americans 2020-2025: 15% for protein, 55% for carbohydrates, and 35% for fats [26]. Although the pipeline uses this distribution profile by default, these percentages can be easily adjusted by the user to accommodate specific dietary requirements or preferences. These targets for macronutrients and calories further guide the MILP solver in the optimization process.

The primary objective of MILP solver in the proposed model is to minimize the deviation from the nutritional targets for calories, protein, carbohydrates, and fats prescribed for the patient. To achieve this, a goal programming problem is formulated in a way that the model aims to minimize the sum of surplus and deficit deviations from each nutritional target. This way, the objective function seeks the meal plan that is closest to ideally matching the desired nutritional profile.

The MILP solver was configured to accommodate a range of dietary and structural constraints in order to generate nutritionally balanced and practically feasible meal combinations. The following paragraphs present an overview of the three main categories of implemented constraints.

##### Nutritional Accuracy

The constraints on the nutritional alignment are set to ensure that the total nutrients of produced meal plans, adjusted by surplus and deficit deviation variables, meet the prescribed targets. Apart from the primary nutritional goals, the solver also incorporates strict health-oriented sodium and potassium intake constraints (maximum of 760 mg of sodium and minimum of 1570 mg of potassium per meal) defined according to recommendations from Dietary Guidelines for Americans 2020-2025 [26].

##### Dietary Structure and Variety

Several rules are imposed to enforce structure and dietary variety in the meal plan, thereby reducing the likelihood of random or illogical combinations of food items. Each meal is set to five food categories, with only one item permitted per category to encourage diversity. In addition, the constraints ensure the inclusion of food items from the defined set of mandatory categories (main dish, soup, salad, and drink) to provide a complete meal plan. The balance of the meal structure is also achieved using a heuristic constraint limiting the caloric content of any single food item to no more than 70% of the main dish’s total calories.

##### Practicality

Serving size constraints are employed by the model to standardize portion options across food items. Instead of allowing continuous gram values, a discrete set of eight possible serving sizes is defined: 50g, 100g, 150g, 200g, 250g, 300g, 350g, and 400g. Research shows that although it is essential to eat appropriate portion sizes, most individuals find it difficult to adhere to portion control [27]. This way the discrete set of serving sizes that are easy-to-estimate ensures more practical and user-friendly recommendations which, in turn, can increase the likelihood of adherence. Moreover, this quantization allows the solver to operate within a practical range of serving portions while maintaining flexibility in meal construction. These serving sizes are further constrained by category-specific rules illustrated in Table 3 which reflect realistic consumption patterns [26].

**Table 3.**
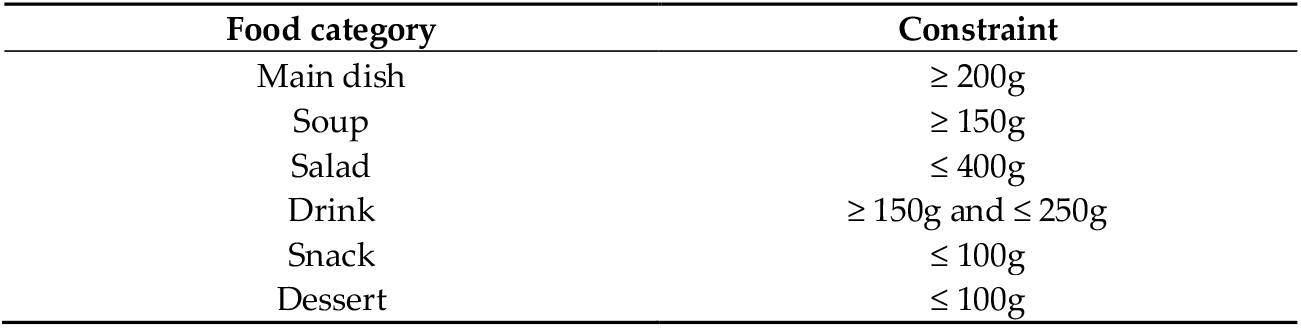
Serving size constraints for each food category posed to the MILP solver.

| Food category | Constraint |
| --- | --- |
| Main dish | $\geq 200\text{g}$ |
| Soup | $\geq 150\text{g}$ |
| Salad | $\leq 400\text{g}$ |
| Drink | $\geq 150\text{g}$ and $\leq 250\text{g}$ |
| Snack | $\leq 100\text{g}$ |
| Dessert | $\leq 100\text{g}$ |

The mathematical formulation of the MILP solver is defined as follows:

##### Sets

- *F*: set of all available food items after LLM filtering
- *N*_*obj*_: set of nutrients included in the objective function, *N*_*obj*_ = {*Calories, Proteins, Carbs, Fats*}.
- *N*_*const*_: set of nutrients managed by hard constraints, *N*_*const*_ = {*Sodium, Potassium*}.
- *C*: set of all food categories, *C* = {*Main dish, Soup, Drink, Salad, Snack, Dessert*}.
- *C*_*mand*_: set of mandatory food categories, *C* = {*Main dish, Soup, Drink, Salad*}.
- *S*: set of discrete serving sizes in grams, *S* = {50,100,150,200,250,300,350,400}.
- *K*: set of solutions found so far in the iterative process. Calories, Proteuns, Carbs, Fats

##### Parameter

- *a*_*i,j*_ : amount of nutrient *j* ∈ *N* per gram of food *i* ∈ *F*.
- *w*_*j*_: grams to calorie conversion factor for nutrient *j* ∈ *N*, (*w*_*calories*_ = 1, *w*_*proteins*_ =*w*_*carbs*_= 4, *w*_*fats*_= 9).
- *T*_*j*_ : The target caloric value for nutrient *j* ∈ *N*.
- *T*_*sad*_ = 760*mg*: maximum amount of sodium per meal.
- *T*_*pot*_ = 1570*mg*: minimum amount of potassium per meal.
- *F*_*c*_: The subset of foods *F* belonging to category *c* ∈ *C*.
- *S*_*min,c*_, *S*_*max,c*_: The minimum and maximum serving sizes for category *c* ∈ *C*.
- *M*_*items*_ = 5: maximum number of items allowed in a single meal.

##### Variables

- *x*_*i,s*_ ∈ {0,1}: binary variable, 1 if item *i* ∈ *F* chosen with serving *s* ∈ *S*.
- *y*_*i*_ ∈ {0,1}: binary variable, 1 if item *i* ∈ *F* item chosen at any serving.
- 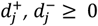: surplus and deficit deviations for *j* ∈ *N*_*obj*_.

##### Objective Function

Minimize

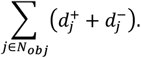

##### Constraints

**Nutrient Balance:** for each nutrient *j* ∈ *N*_*obj*_, the total calories must equal the target, adjusted by deviation variables.

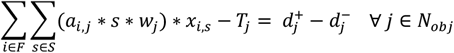

**Sodium & Potassium:** hard nutritional constraints imposed on sodium and potassium.

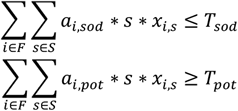

**Serving Size Uniqueness:** each food item can only have one serving size.

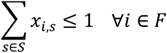

**Linking x and y:** the variable *y*_*i*_ is activated if any serving size is chosen for food *i* ∈ *F*.

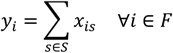

**Total Items:** the total number of distinct food items in the meal is limited

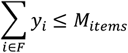

**Category at-most-one:** at most one item can be selected from each category.

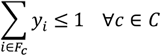

**Mandatory Categories:** the meal must contain at least one item from each mandatory category.

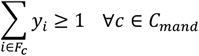

**Category-Specific Serving Sizes:** for each food the chosen serving size must be within the allowed range.

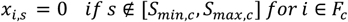

**Main Dish Caloric Dominance:** the calories of any single non-main dish item must not exceed 70% of the total calories of all main dish items.

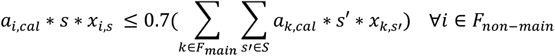

**Iterative Solution Exclusion:** to generate distinct solutions for each new run, a constraint is added that forbids exact sets of food items from all previously found solutions.

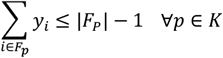

#### 2.1.4 Final Selection by LLM

The next stage of the proposed pipeline involves qualitative selection handled by the LLM. The 10 best meal plans offered by the MILP solver are passed back to the GPT-4o model which, in turn, analyzes these options in the context of the given patient profile and selects the single most appropriate meal plan. The LLM is also tasked with providing a detailed rationale for its choice. This approach leverages both the optimization strength of MILP solver and reasoning capabilities of the LLM, enabling solutions that align closely with target calories and account for user-specific logic and preferences. As for the initial part, the LLM is guided by a structured prompt given below:

“You are an expert dietitian and meal planning assistant. Your task is to analyze a list of meal plan options and select the single best one for a user based on their profile and preferences. Then, you must provide a clear, encouraging, and detailed rationale for your choice.”

For the user prompt, the patient’s nutritional targets and the formatted list of 10 optimal meal plans are included. The LLM is instructed to output the index of the selected meal combination as well as the detailed rationale for its choice. The configuration of system parameters remained the same, ensuring consistency in the model responses.

#### 2.1.5 AI Image Generation for Visual Representation

The final step of our pipeline involves AI-based generation of a photorealistic image of the recommended meal plan, aimed to improve the user experience and promote dietary adherence. For this task, Google’s Vertex AI Imagen v0.6 text-to-mage diffusion model is utilized. After the final meal plan is selected by LLM, the system constructs a descriptive text prompt incorporating the list of food items present in the chosen plan and sends it directly to the Imagen API for the extraction of visual representation of the meal.

### 2.2. Comparative Models

To assess the performance of the proposed hybrid MILP+LLM model, two following standalone models were developed for comparison.

#### Standalone MILP model

This model utilizes only the MILP solver for meal plan generation. Unlike in the hybrid model, the solver receives the entire, unfiltered dataset with food items. The same nutritional targets, objective function as well as the set of constraints as in the hybrid model are used. The solver’s primary objective is to find the single most mathematically optimal solution based on the patient-specific nutritional targets without any prior personalization or further qualitative selection.

#### Standalone LLM model

This model uses solely the LLM (GPT-4o) to produce a meal combination. The LLM is given a comprehensive and well-structured prompt containing the complete patient profile. The prompt explicitly instructs the model to first calculate the user-specific caloric target. Crucially, the entire food dataset is also provided to the LLM as a part of the prompt, strictly limiting the model to the available options only. When tasked with generating the meal plan, the LLM is instructed to adhere to a set of constraints identical to those imposed to the MILP solver in the hybrid model. This way, the standalone LLM model aims to identify the most appropriate meal combination within a single generation step and without any external mathematical solver.

### 2.3. Evaluation Methodology

To evaluate and compare the three models (MILP+LLM, MILP, and LLM), synthesized patient profiles were utilized, directly sourced from the study by Adilmetova et al. [4]. To showcase the proposed model’s ability to generate meal plan recommendations, five cases were selected from the available profiles, each involving individuals with obesity, diabetes, or weight-management objectives, which require careful consideration and understanding of the context. A total of 15 recommendations (5 patient profiles × 3 models) were extracted and further utilized for comparative evaluation of functionality of the proposed method.

#### 2.3.1. Evaluation Rubric

A comprehensive evaluation rubric was introduced to assess four metrics essential for the effectiveness and quality of personalized nutritional advice: Nutrient Accuracy, Personalization, Practicality and Variety.

**Nutrient Accuracy:** assesses the extent to which the meal plan aligns with desired nutritional targets, including calories, protein, carbohydrates, and fats as well as maximum or minimum requirements for sodium and potassium.

**Personalization:** approximates the degree to which the response is tailored to the specific needs, characteristics, and context of the individual, rather than providing generic advice, or, essentially, relevance to the person’s health status, dietary preferences, cultural background, and stated goals.

**Practicality :** determines the feasibility, realism, and real-world applicability of the advice provided, including whether the information can be readily applied by the individual in their daily life; considers factors such as resource availability, time constraints, and ease of implementation.

**Variety:** measures the diversity of food groups available in the meal plan; evaluates balance across fruits, grains, vegetables, protein sources, and dairy.

#### 2.3.2. Evaluation of Responses

The rubric is based on a 5-point Likert scale used for all models, with each of the four evaluation criteria being rated from 1 (very poor) to 5 (excellent) (Appendix 2). This scaling system was selected for several reasons that set up the environment for precise systematic analysis. A 5-point scale provides a midpoint (neutral/acceptable), crucial for nuanced assessments and adequate expressions of opinions behind the decisions, which is not available in scales with four or fewer points [28]. Research findings suggest that when comparing 4-point and 5-point scales, versions of the latter demonstrate higher internal consistency and better validity [28]. In other words, they produce more consistent responses and can better distinguish between multiple layers of quality. Furthermore, 5-point scales are often adopted in other works focused on similar evaluation tasks [27, 29-33]. The chosen method also tends to minimize the amount of missing data and is usually preferred by the responders [28].

#### 2.3.3. Evaluation Team

The generated meal plans were assessed by a team of five evaluators, including medical doctors, public health care specialists, and dietitians. Each evaluator reviewed all 15 meal recommendations from the three models, using the above-mentioned evaluation rubric based on nutrient accuracy, personalization, practicality, and variety.

#### 2.3.4. Descriptive and Statistical Analyses

Descriptive and statistical analyses were conducted using Stata 18. Initially, descriptive statistics including the mean were calculated for each of the four criteria to summarize the performances of the three models.

Non-parametric tests were used for inferential analysis because the 5-point Likert scale data is ordinal in nature. To ascertain whether there were statistically significant variations in the assessment scores between the three models, a Kruskal-Wallis H test was conducted for each criterion. A Dunn’s post-hoc test for pairwise comparisons with a Bonferroni correction for multiple tests was carried out after each Kruskal-Wallis test result. This allowed for pinpointing important distinctions between all of the model pairs (MILP vs LLM, MILP+LLM vs LLM, and MILP+LLM vs MILP). A p-value of 0.05 was regarded as a level of significance for the statistical tests.

## 3. Results

### 3.1. Component-Level Performance of the Hybrid MILP+LLM Model

Before providing the comparative evaluation of the three models, this section assesses the performance of each component within the proposed hybrid MILP+LLM model to demonstrate its internal efficacy and efficiency. A complete sample of the output generated by the pipeline for a single patient profile is provided in Appendix 3 for further reference.

The first stage utilizes an LLM to filter the main dataset by separating the food classes that are considered permissible for a given user profile. The effectiveness of this module directly impacts the extent of personalization and time complexity of the MILP solver. On average, the LLM identified and removed 86% of the food items from the original dataset, successfully eliminating those contraindicated by health conditions, known allergens, or explicitly stated user preferences. Notably, initial filtering conducted by the LLM contributed to a considerable reduction in the time complexity of the whole model discussed later in this section. A manual audit of this process confirmed that the LLM-filtering step reliably retained a context-appropriate set of food classes, thereby enhancing the relevance of the inputs and the efficiency of the subsequent optimization stage.

Based on the filtered dataset from the LLM, the next component of the pipeline employs the MILP solver, enabling the meal plan recommendations to be not only personalized, but also nutritionally precise. A critical observation of this validation is that the solver was able to successfully find a set of feasible solutions in all five cases. Therefore, it can be concluded that the LLM-filtering module, while significantly restrictive, never narrowed down the solution space to the point where it became impossible to meet the target nutritional values. For every patient profile, the MILP solver consistently identified a set of 10 distinct optimal solutions of meals that strictly adhered to the user’s calculated caloric and macronutrient targets. The combinations of food within the ten solutions were constantly reasonable and practical, which confirms the component’s high reliability and accuracy within the pipeline.

A primary advantage of the proposed hybrid model is its computational efficiency. By significantly reducing the solution space before optimization, the LLM-filtering stage allows the proposed model to determine optimal solutions much faster when compared to the standalone MILP model. As shown in Table 4, the standalone LLM model achieved the shortest computation time on average, followed by the hybrid MILP+LLM model, while the standalone MILP required the longest time across all patients. The hybrid model outperformed the standalone LLM in two of the five patients (patients 31 and 35). It is worth noting that the computation time for the proposed model includes the complex task of finding 10 distinct optimal solutions, whereas the standalone MILP model’s time reflects identifying only a single best meal plan. Since the hybrid model was faster than the standalone MILP solver in all cases, it can be concluded that the LLM pre-filtering module has a significant impact on reducing the computational load.

**Table 4.** Meal plan computation times (in seconds) of three models across all five synthetic patient profiles.

| Dataset | Patient 7 | Patient 31 | Patient 35 | Patient 40 | Patient 46 |
| --- | --- | --- | --- | --- | --- |
| MILP | 119.84 | 1013.78 | 335.89 | 2831.67 | 2709.71 |
| LLM | 23.95 | 48.13 | 38.49 | 43.91 | 47.41 |
| MILP+LLM | 36.04 | 33.08 | 28.90 | 90.34 | 749.75 |

The final LLM component serves as a qualitative reasoning layer aimed to select the ultimate meal plan from the top 10 solutions provided by the MILP solver. This module succeeded in its function across all patient profiles. Notably, the LLM did not always choose the lowest cost function value solution. Instead, it often prioritized plans that better aligned with a patient’s unique needs, such as lifestyle constraints or cultural food preferences, which are difficult to encode in a mathematical model. As exemplified by the sample provided in Appendix 3, the accompanying rationales consistently translated the output data into personalized dietary advice by linking specific food choices to the patients’ clinical needs and addressing the patient directly, which kept the rationale well-structured, respectful, and aware of the user context.

The pipeline’s final component generates a visual representation of the recommended meal plan using Vertex AI’s Imagen model. This feature is designed to aid user motivation and adherence to the diet by providing a clear, appetizing picture of the meal. In general, the generated images were photorealistic and correlated to the required portion sizes.

### 3.2. Comparative Evaluation of Model Outputs

Overall, the standalone LLM model consistently received the lowest scores across most of the evaluation metrics. The highest average for the LLM model is 3.67 in Practicality, where all three models are relatively close. In contrast, both the hybrid MILP+LLM model and the purely optimization-based MILP model achieved considerably stronger performance across the evaluation criteria. The hybrid MILP+LLM model was found to consistently produce stable and high-quality outputs with its average scores remaining consistently above 3.6 points for all evaluation criteria. On the other hand, the standalone MILP approach showed greater range with its scores varying from a high of 4.93 in Nutrient Accuracy to a low of 3.43 in Personalization. The MILP model achieved the highest scores for Nutrient Accuracy (4.93) and Variety (4.10), whereas the combined approach MILP+LLM delivered the top scores of 3.99, 3.96, and 3.81 for Practicality, Nutrient Accuracy, and Personalization, respectively, indicating its strength in producing feasible, accurate and tailored meal plans.

### 3.3 Statistical Significance Analysis

The Kruskal–Wallis tests (see Table 5) revealed statistically significant differences across models for nutrient accuracy (χ^2^ = 158.994, p < 0.0001), personalization (χ^2^ = 34.670, p < 0.0001), and variety (χ^2^ = 30.962, p < 0.0001). A smaller but still significant difference was observed for practicality (χ^2^ = 6.122, p = 0.0468). These findings indicate that evaluators perceived meaningful variations in model performance across multiple evaluation domains.

**Table 5.** Kruskal–Wallis rank test summary.

| Criterion | $\chi^2$ (df = 2) | p-value |
| --- | --- | --- |
| Nutrient Accuracy | 158.994 | 0.0001 |
| Personalization | 34.670 | 0.0001 |
| Practicality | 6.122 | 0.0468 |
| Variety | 30.962 | 0.0001 |

Pairwise post-hoc Dunn’s tests with Bonferroni adjustment in Table 6 further clarified these differences. For accuracy, both MILP and MILP+LLM received significantly higher ratings than LLM alone (p < 0.0001). Moreover, MILP was rated significantly higher than MILP+LLM (p < 0.0001), suggesting that evaluators perceived the linear programming approach to be the most accurate.

**Table 6.**
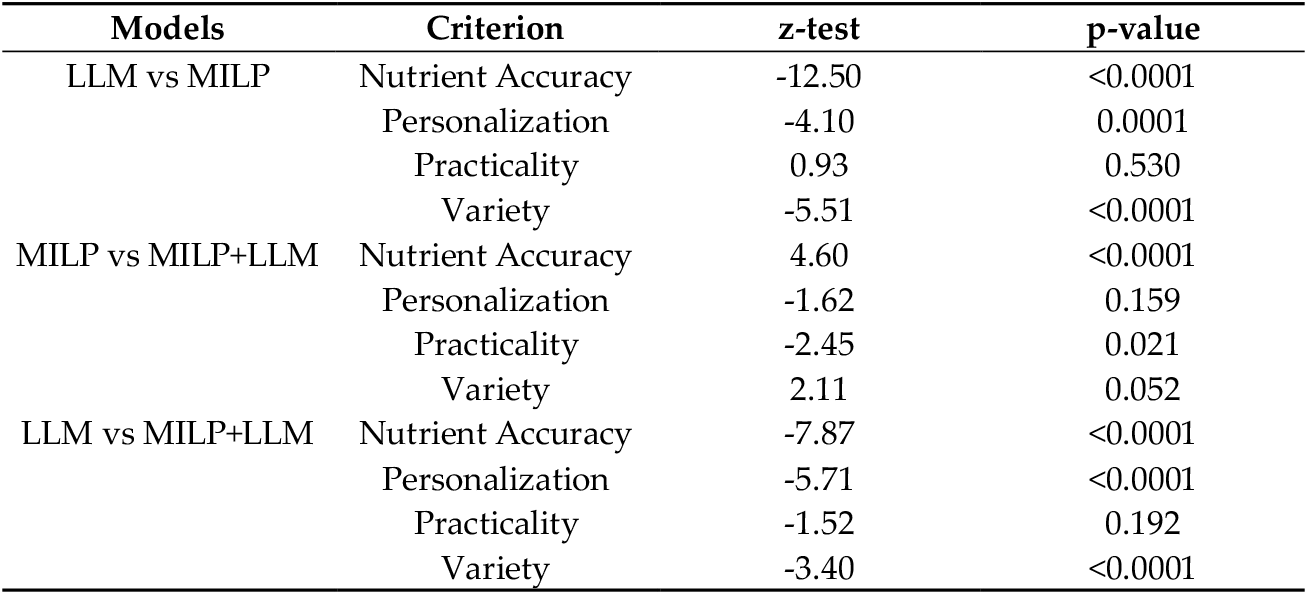
Post hoc Dunn’s test results.

| Models | Criterion | z-test | p-value |
| --- | --- | --- | --- |
| LLM vs MILP | Nutrient Accuracy | -12.50 | <0.0001 |
|  | Personalization | -4.10 | 0.0001 |
|  | Practicality | 0.93 | 0.530 |
|  | Variety | -5.51 | <0.0001 |
| MILP vs MILP+LLM | Nutrient Accuracy | 4.60 | <0.0001 |
|  | Personalization | -1.62 | 0.159 |
|  | Practicality | -2.45 | 0.021 |
|  | Variety | 2.11 | 0.052 |
| LLM vs MILP+LLM | Nutrient Accuracy | -7.87 | <0.0001 |
|  | Personalization | -5.71 | <0.0001 |
|  | Practicality | -1.52 | 0.192 |
|  | Variety | -3.40 | <0.0001 |

For personalization, MILP and MILP+LLM significantly outperformed LLM (p ≤ 0.0001). Moreover, MILP + LLM achieved a slightly higher mean score (3.81) than standalone MILP (3.43), indicating that integrating an LLM into MILP further enhanced perceived personalization.

For practicality, no significant differences were observed between MILP and LLM (p = 0.530) or MILP+LLM and LLM (p = 0.192). However, MILP was rated significantly less practical than MILP+LLM (p = 0.021), suggesting that integration of the two approaches may have improved practicality.

For variety, both MILP and MILP+LLM were rated significantly higher than LLM alone (p < 0.0001). A trend toward higher ratings for MILP compared with MILP+LLM was observed (p = 0.052), although it did not reach statistical significance.

## 4. Discussion

The aim of this study was to introduce and evaluate a hybrid model that integrates a MILP solver together with an LLM to generate personalized meal plans. The findings suggest that the proposed MILP+LLM approach consistently outperforms a standalone LLM model and excels in personalization and practicality when compared to LLM-only model.

Based on several statistical analyses, the MILP+LLM model consistently scored high points from the rubric, averaging more than 3.6 across each of the four evaluation metrics. The stability in the performance of the proposed method is a sign that both the LLM and the numeric precision behind the MILP solver complement each other well. It can also be inferred that the initial filtering of the food dataset by the LLM does narrow down the choices to a subset that is tailored to the user’s medical profile for the MILP solver. The findings also suggest that an additional layer of LLM that selects the most appropriate meal plan from the 10 optimal solutions provided by the MILP solver further expands the performance of the MILP+LLM model across the evaluation metrics. It can be concluded that the proposed approach truly leverages the benefits of quantitative optimization with qualitative and patient-specific factors.

The results for Nutrient Accuracy mostly aligned with theoretical expectations. As anticipated, the standalone MILP solver achieved an almost perfect score of 4.93, confirming the strong performance of a mathematical solver in constrained numerical optimization tasks. Conversely, the standalone LLM model scored a dramatically low 1.54. This finding suggests that although LLMs possess advanced reasoning capabilities, they are unreliable for tasks involving strict mathematical adherence where precision is extremely important. This observation reflects that of Bergling et al. [13] who also reported the frequent failure of LLMs to perform accurate numerical calculations of nutrient values, a considerable limitation in the context of personalized meal planning. Interestingly, the proposed MILP+LLM model, while scoring quite high (3.96), was slightly outperformed by the sole MILP solver. This is likely due to the initial filtering step handled by the LLM which, despite its benefit for personalization, reduces the total number of available food items, thereby slightly constraining the integrated solver’s ability to find perfectly nutritionally optimized solution.

In the Personalization metric, the proposed hybrid model achieved the highest score of 3.81. It makes it evident that the combined approach is crucial in generating nutritional advice that fits the user’s unique needs and preferences. This discovery is aligned with the rising recognition that more tools, including LLMs, should be applied in further development of providing well-rounded meal plans for individuals seeking dietary advice [34]. Surprisingly, the standalone LLM model scored the lowest of the three models at 2.60. A possible explanation for this might be the cognitive load of the prompt that required the LLM to act as a dietitian, analyze a large dataset, and at the same time adhere to the set of numerical constraints. This variety and load in tasks may have negatively affected the model’s personalization capabilities. Another unanticipated result was the standalone MILP model’s respectable score of 3.43. This finding is contrary to existing research which suggests that sole MILP solvers often fail to personalize and adapt to important individual dietary factors such as health conditions, cultural norms and taste preferences [35]. It turns out that accurately meeting a patient’s specific nutritional targets is itself a crucial aspect of effective personalization.

For the Practicality criterion, the differences between scores achieved by models were less pronounced, showing that all three approaches managed to deliver reasonable and practical meal suggestions. The hybrid MILP+LLM system once again achieved the highest score (3.99). This advantage can be attributed to the final qualitative selection step conducted by the LLM. Provided with a list of 10 nutritionally optimal plans, the LLM effectively avoids less coherent meal combinations (e.g. including inappropriate food pairings), ultimately selecting the most practical one that could be easily implemented by the user in their daily life.

In terms of Variety, the standalone MILP model outperformed the other two models and scored 4.10. This result highlights an extremely important architectural trade-off. The MILP solver utilized the whole unfiltered dataset which provided the model with the maximum possible number of food items to create diverse combinations. In contrast, due to the initial filtering step, for the hybrid MILP+LLM model the dataset was narrowed down significantly, reducing the potential for sheer variety. However, this trade-off emphasizes a considerable advantage of the proposed hybrid model: computation time. During the meal generation process for each of the models, it was observed that it took substantially longer for the standalone MILP solver to find the optimal solution due to larger search space. This way, the hybrid MILP+LLM system’s pre-filtering step not only facilitates increased personalization but also makes the optimization process far more computationally efficient, highlighting an advantage of the proposed solution.

The method proposed in this study tackles one of the challenges that AI-driven health recommendation services often face – user trust. On their own, LLMs are “black box” operators, which interferes with establishing open, reliable, and accessible connections with the individuals who may be hesitant to follow dietary advice without evidence [32]. The hybrid MILP+LLM model surpasses the sole LLM approach in this regard by offering interpretability behind its calculations, which ensure that nutritional targets are met. This layer for careful personalization and detailed explanation could potentially increase the level of user confidence in the provided meal plans, yielding a more accessible and understandable output.

## 5. Limitations

This study has few limitations that should be acknowledged and can be addressed in future work. The proposed solution’s clinical applicability can be significantly enhanced by expanding the food dataset with a more diverse and globally curated food options. Moreover, the current model is designed to optimize a single meal rather than to generate a daily or weekly dietary plan. From that perspective, it does not take into account leftovers from previous meals, cumulative costs, and sufficient micronutrients for long-term dietary goals. Building on the research conducted in this study, future steps will involve expanding the model to generate full day eating plans and incorporating economic and food availability constraints to better serve the needs of users.

## Supporting information

Appendix 2. Evaluation Rubric

Appendix 3. Sample Output

## Author Contributions

Conceptualization, H.A.V. and M.-Y.C.; methodology, R.A., I.N., D.K., H.A.V. and M.-Y.C.; validation, R.A., I.N. and D.K.; formal analysis, R.A., I.N., D.K, B.N. and Z.O.; investigation, R.A., I.N. and D.K.; resources, M.-Y.C. and H.A.V.; data curation, R.A. and I.N.; writing—original draft preparation, R.A., I.N., D.K. and M-Y.C; writing—review and editing, R.A., I.N., D.K., B.N., Z.O., H.A.V. and M.-Y.C.; visualization, R.A and B.N.; supervision, M.-Y.C. and H.A.V.; project administration, M.-Y.C. and H.A.V.; funding acquisition, M.-Y.C. and H.A.V. All authors have read and agreed to the published version of the manuscript.

## Funding

This research is funded by the Science Committee of the Ministry of Science and Higher Education of the Republic of Kazakhstan (Grant No. AP23485288) and Nazarbayev University, under the Faculty Development Competitive Research Grant Program (Grant No. 201223FD2603).

## Institutional Review Board Statement

The present study utilized five mock patient profiles to generate AI-based dietary recommendations for individuals with diabetes or obesity. The study does not contain real patient data, ethical approval was hence unnecessary from the Ethics Committee.

## Data Availability Statement

The model outputs generated in this study will be made publicly and freely available without restriction after the publication of the manuscript.

## Acknowledgments

The authors acknowledge the evaluation team members, including Ilvira Ibraimova, Nurgul Makhmetova, Aizhan Meyerbekova, Zhannur Toreber, and Dina Sabyr, for their careful participation and assistance in the assessment process.

## Conflicts of Interest

The authors declare no conflicts of interest.

## Abbreviations

The following abbreviations are used in this manuscript:

BMR: Basal metabolic rate
LLM: Large language model
LP: Linear programming
MILP: Mixed integer linear programming
MPG: Meal plan generator
PN: Personalized nutrition
RAG: Retrieval-augmented generation
TDEE: Total daily energy expenditure

## Appendix

### 1. Sample Patient Profile [4]

Case 35: Obesity. Dyslipidemia. Name: Tamara Gender: Female Age: 49 Nationality: Russian Location: Pavlodar, Kazakhstan Family Information: Married with two daughters who are students. Responsible for caring for her mother with dementia. Occupation: Full-time call center operator. Presented complaints: Weight gain of 15 kg, high blood pressure up to 160/90 mmHg. Diagnosis: Obesity. Dyslipidemia. Arterial hypertension. Medical History: There is no history of weight problems before marriage. Weight gain of over 15 kg during each pregnancy, unable to lose weight while breastfeeding. She also has history of arterial hypertension for 3 years. Anthropometry, Body Composition, and Functional: Weight: 106 kg Height: 1.60 m, BMI 41,4 kg/m2 Physical examination: BP 165/90 mmHg, HR 65 bpm, SatO2 97%. Auscultation of the lungs reveals vesicular breathing, no wheezing. The heart sounds is clear, there are no murmurs. Physical examination revealed abdominal enlargement due to subcutaneous fat. Diagnostic work-up: LDL Cholesterol: 4.1 mmol/L, HDL Cholesterol: 0.9 mmol/L. Management: Eating a healthy, balanced diet and regular physical activity. Control BP. Valsartan 80 mg twice daily to control hypertension, Atorvastatin 20 mg to control cholesterol levels, Semaglutide (Ozempic) injection weekly for weight control. Diet: Tried various commercial diets but it was not effective. Environmental, Behavioral, and Social Factors: Tamara works full time in office. She takes care her mother who has dementia Additional Information: She is concerned about her weight and its impact on her health. She is under increased risk of stroke and heart problems due to high blood pressure and cholesterol levels. She is uncertain about bariatric surgery due to caregiving responsibilities for her mother and work commitments. She worries about traveling to another city for surgery and potential complications of anesthesia. Her family is concerned about the risks of stroke and heart disease if her condition is left untreated.

### 2. Evaluation Rubric (attached in PDF)

### 3. Sample Output (attached in PDF)

## Disclaimer/Publisher’s Note

The statements, opinions and data contained in all publications are solely those of the individual author(s) and contributor(s) and not of MDPI and/or the editor(s). MDPI and/or the editor(s) disclaim responsibility for any injury to people or property resulting from any ideas, methods, instructions or products referred to in the content.

## Notes

### Competing Interest Statement

The authors have declared no competing interest.

