## Appendix 2. Evaluation Rubric for "ChatGPT with Mixed-Integer Linear Programming for Precision Nutrition Recommendations"

| Criteria | 1 – Very Poor | 2 – Poor | 3 – Acceptable | 4 – Good | 5 - Excellent |
|---|---|---|---|---|---|
| **Nutrient Accuracy**<br><br>How well the meal plan matches nutrient targets (macros and micros). | At least one macronutrient is off by more than 35%.<br><br>If sodium is above its limit, it is more than 50% over.<br><br>If potassium is below its minimum, it is more than 50% under. | At least one macronutrient is off by 20% to 35%.<br><br>If sodium is above its limit, it is between 26% and 50% over.<br><br>If potassium is below its minimum, it is between 26% and 50% under. | All macronutrients are within 20% of their targets.<br><br>If sodium is above its limit, it is between 11% and 25% over.<br><br>If potassium is below its minimum, it is between 11% and 25% under. | Most macronutrients are within 10% of their targets (none more than 15% off).<br><br>If sodium is above its limit, it is no more than 10% over.<br><br>If potassium is below its minimum, it is no more than 10% under. | All macronutrients are within 5% of their targets.<br><br>Sodium does not exceed its limit.<br><br>Potassium meets or exceeds its minimum. |
| **Personalization**<br><br>How well the meal plan fits the patient's individual profile (preferences, restrictions, clinical needs). | The meal plan ignores the patient profile entirely or includes foods that could be harmful or contraindicated. | Limited adaptation to the patient — multiple foods are unsuitable or clearly not personalized. | Generally appropriate, but several food items seem generic or not clearly tailored to the patient profile. | Mostly adapted, with only minor mismatches (for example, one less suitable item).<br><br>No serious contraindications. | Fully adapted to the patient's medical needs, preferences, restrictions, and cultural considerations.<br><br>No inappropriate or contraindicated foods. |
| **Practicality**<br><br>How realistic and feasible the meal plan is for everyday implementation. | The meal plan is largely impractical — requires special ingredients, extensive effort, or very high cost. | Several items are hard to source, expensive, or unrealistic in portion size. | Some impractical elements, such as moderate cost, preparation difficulty, or uncommon ingredients. | Mostly practical with only minor cost, availability, or preparation concerns. | All food is affordable, widely available, and simple to prepare.<br><br>Portion sizes are reasonable for everyday use. |

| | | | | | |
|---|---|---|---|---|---|
| **Variety**<br><br>How well the meal plan includes diverse food groups (fruits, grains, vegetables, protein, dairy). | Includes foods from only one group, or multiple meals lack variety (e.g., mainly grains). | Covers only two food groups; large gaps in diversity (e.g., no vegetables or dairy). | Includes at least three groups, but balance is limited (e.g., vegetables present but no fruit). | Includes four groups with reasonable balance across meals. | All five groups are present, well balanced across meals, and no group is overemphasized. |
