## Appendix 3. Sample Output for "ChatGPT with Mixed-Integer Linear Programming for Precision Nutrition Recommendations"

### Nutritional Meal Planner

##### Tell us about yourself and your meal preferences:

Please describe your situation, health conditions, dietary restrictions, goals, and preferences.
**Examples:**
- "I'm a 45-year-old male with type 2 diabetes. I need a low-carb lunch and dislike fish."
- "I'm underweight and lactose intolerant. I want a high-calorie, high-protein meal."
- "I'm an athlete looking for a vegetarian post-workout recovery meal."

```
Obesity. Dyslipidemia. Name: Tamara Gender: Female Age: 49 Nationality: Russian Location: Pavlodar, Kazakhstan Family Information: Married with two daughters who are students. Responsible for caring for her mother with dementia. Occupation: Full-time call center operator. Presented complaints: Weight gain of 15 kg, high blood pressure up to 160/90 mmHg. Diagnosis: Obesity.  Dyslipidemia. Arterial hypertension. Medical History: There is no history of weight problems before marriage. Weight gain of over 15 kg during each pregnancy, unable to lose weight while breastfeeding. She also has history of arterial hypertension for 3 years. Anthropometry, Body Composition, and Functional: Weight: 106 kg Height: 1.60 m, BMI 41,4 kg/m2 Physical examination:  BP  165/90 mmHg, HR 65 bpm, SatO2 97%. Auscultation of the lungs reveals vesicular breathing, no wheezing. The heart sounds is clear, there are no murmurs. Physical examination revealed abdominal enlargement due to subcutaneous fat. Diagnostic work-up:  LDL Cholesterol: 4.1 mmol/L, HDL Cholesterol: 0.9 mmol/L. Management: Eating a healthy, balanced diet and regular physical activity. Control BP. Valsartan 80 mg twice daily to control hypertension, Atorvastatin 20 mg to control cholesterol levels,  Semaglutide (Ozempic) injection weekly for weight control. Diet: Tried various commercial diets but it was not effective. Environmental, Behavioral, and Social Factors: Tamara works full time in office. She takes care her mother who has dementia Additional Information: She is concerned about her weight and its impact on her health. She is under increased risk of stroke and heart problems due to high blood pressure and cholesterol levels. She is uncertain about bariatric surgery due to caregiving responsibilities for her mother and work commitments. She worries about traveling to another city for surgery and potential complications of anesthesia. Her family is concerned about the risks of stroke and heart disease if her condition is left untreated.
```

**Age:**

49

**Weight (kg):**

106

**Height (cm):**

160

**Sex:**

Female

**Activity Factor:**

| Level | Description | Factor |
|---|---|---|
| Sedentary | Little or no exercise, desk job | 1.2 |
| Lightly active | Light exercise/sports 1–3 days/week | 1.375 |
| Moderately active | Moderate exercise/sports 3–5 days/week | 1.55 |
| Very active | Hard exercise 6–7 days/week | 1.725 |
| Extra active | Very intense training, physical job, or 2× daily workouts | 1.9 |

Sedentary (1.2)

**Protein (% of Calories): 15%**

**Carbs (% of Calories): 50%**

**Fats (% of Calories): 35%**

Generate Meal Plan

**Your calculated calories for this meal:** 662 kcal
(Based on your BMR and activity, divided by 3 for one meal)

##### Step 1: Foods Selected by the LLM Filter

| | | | | |
|---|---|---|---|---|
| oatmeal porridge | fresh vegetables | fresh salad | cottage cheese | kefir |
| greek salad | beef | chicken | salmon with rice | vegetable shashluk |
| buckwheat | lentil soup | beet salad | vegetables stew | water |
| steamed buckwheat | broccoli | hummus | vegetarian stew | vegetable soup |
| minestrone soup | veggie burger | salmon cooked | tuna salad | banana |
| blueberries | apple | raisins | apricot dried | edamame cooked |
| chickpeas cooked | greek yogurt | tap water | black tea | herb tea |
| almonds raw | walnut | vegetable salad | arugula | coleslaw |

**Total Foods Selected:** 40

##### Step 2: Top 10 Solutions from MILP Solver

**Solution 1 (Score: 35.39)**

- fresh vegetables: 50g
- salmon with rice: 250g
- minestrone soup: 150g
- black tea: 250g

Calories: 677, Proteins: 27.8g, Carbs: 84.0g, Fats: 25.3g

**Solution 2 (Score: 35.39)**

- fresh salad: 50g
- salmon with rice: 250g
- minestrone soup: 150g
- black tea: 250g

Calories: 677, Proteins: 27.8g, Carbs: 84.0g, Fats: 25.3g

**Solution 3 (Score: 37.70)**

- salmon with rice: 250g
- minestrone soup: 150g
- black tea: 250g
- walnut: 100g
- arugula: 100g

Calories: 666, Proteins: 28.7g, Carbs: 83.6g, Fats: 24.1g

**Solution 4 (Score: 47.13)**

- vegetable shashluk: 400g
- vegetable soup: 150g
- raisins: 50g
- black tea: 250g
- vegetable salad: 350g

Calories: 673, Proteins: 22.5g, Carbs: 81.6g, Fats: 28.2g

**Solution 5 (Score: 48.18)**

- vegetable shashluk: 400g
- vegetable soup: 150g
- raisins: 50g
- tap water: 250g
- vegetable salad: 400g

Calories: 679, Proteins: 23.4g, Carbs: 82.0g, Fats: 28.2g

**Solution 6 (Score: 48.18)**

- vegetable shashluk: 400g
- water: 200g
- vegetable soup: 150g
- raisins: 50g
- vegetable salad: 400g

Calories: 679, Proteins: 23.4g, Carbs: 82.0g, Fats: 28.2g

**Solution 7 (Score: 48.78)**

- vegetable shashluk: 400g
- vegetable soup: 150g
- raisins: 50g
- herb tea: 150g
- vegetable salad: 400g

Calories: 682, Proteins: 23.4g, Carbs: 82.6g, Fats: 28.2g

**Solution 8 (Score: 49.13)**

- vegetable shashluk: 400g
- vegetable soup: 150g
- apple: 50g
- black tea: 250g
- vegetable salad: 350g

Calories: 675, Proteins: 22.3g, Carbs: 82.2g, Fats: 28.3g

**Solution 9 (Score: 49.35)**

- salmon with rice: 250g
- minestrone soup: 150g
- black tea: 200g
- walnut: 100g
- vegetable salad: 50g

Calories: 671, Proteins: 29.0g, Carbs: 84.7g, Fats: 24.1g

**Solution 10 (Score: 49.77)**

- vegetable shashluk: 400g
- vegetable soup: 150g
- blueberries: 50g
- black tea: 250g
- vegetable salad: 400g

Calories: 681, Proteins: 23.4g, Carbs: 82.6g, Fats: 28.5g

---

##### Your Personal Meal Rationale

Dear Tamara, I have selected Option 9 as the best meal plan for you. This option provides a balanced mix of nutrients that align well with your dietary goals and health needs. The meal includes salmon with rice, which is an excellent source of lean protein and omega-3 fatty acids, beneficial for heart health and managing dyslipidemia. The inclusion of minestrone soup and a vegetable salad ensures you receive a variety of vegetables, contributing to your fiber intake and helping with satiety, which is important for weight management. The walnuts add healthy fats, which are crucial for maintaining a balanced diet and can help improve your HDL cholesterol levels. The total calorie count of 671 kcal is very close to your target of 662 kcal, and the macronutrient distribution is well-aligned with your goals of 15%

> protein, 50% carbs, and 35% fats. This meal plan is not only nutritious but also diverse, making it enjoyable and sustainable. Remember, small consistent changes can lead to significant health improvements over time. Keep up the great work on your journey to better health!

#### Your Recommended Meal Plan

| Item | Serving (g) |
|---|---|
| salmon with rice | 250g |
| minestrone soup | 150g |
| black tea | 200g |
| walnut | 100g |
| vegetable salad | 50g |

##### Nutrient Summary

**Calories**
**Actual: 671 kcal**
Target: 662 kcal

**Proteins**
**Actual: 116 kcal**
Target: 99 kcal
Grams: 29.0g

**Carbs**
**Actual: 339 kcal**
Target: 331 kcal
Grams: 84.7g

**Fats**
**Actual: 216 kcal**
Target: 232 kcal
Grams: 24.1g

**Sodium**
**Actual: 437 mg**
Limit: ≤ 760 mg

**Potassium**
**Actual: 1585 mg**
Minimum: ≥ 1570 mg

**Visual Approximation of Your Meal:**

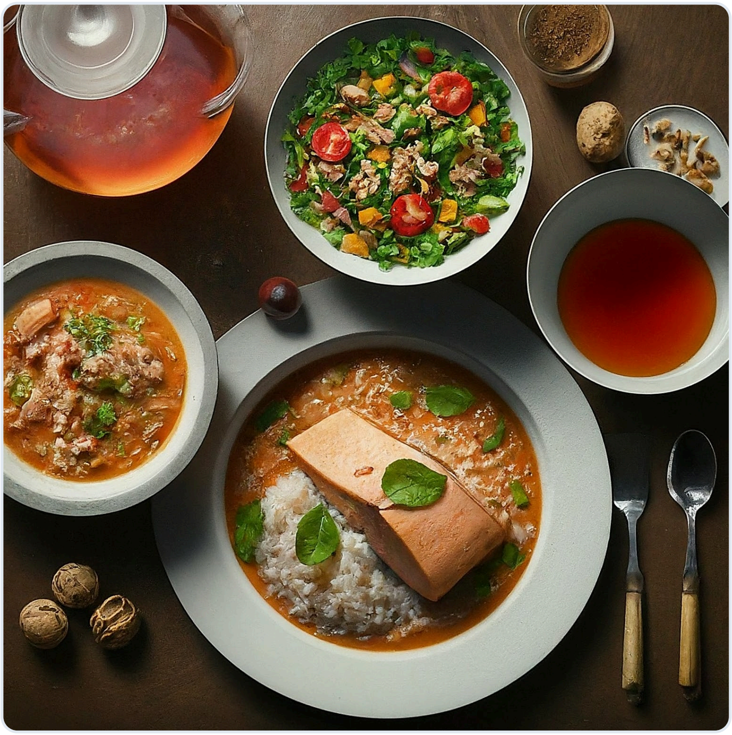

**Meal generated in:** 28.90 seconds
